# COVID-19 in pregnancy – characteristics and outcomes of pregnant women admitted to hospital because of SARS-CoV-2 infection in the Nordic countries

**DOI:** 10.1101/2021.02.05.21250672

**Authors:** Hilde Engjom, Anna JM Aabakke, Kari Klungsøyr, Teresia Svanvik, Outi Äyräs, Eva Jonasdottir, Lars Thurn, Elin Jones, Karin Pettersson, Lill Trine Nyfløt, Iqbal Al-Zirqi, Siri Vangen, Pétur B. Júlíusson, Karin Källén, Mika Gissler, Lone Krebs

**Author notes:** **Corresponding author**: Hilde Engjom, The Norwegian Institute of Public Health, Bergen, Norway and Haukeland University Hospital, Bergen, Norway., or. Shared first authorship. **Conflicts of interest** None of the authors have stated any conflict of interest and all authors have completed the IJME form. **Funding** information The clinical trial management software EasyTrial, used for data management in Denmark, was provided for free for this study as the company offered free access to the software to COVID-19 research projects. The Nordic Federation of Societies of Obstetrics and Gynecology (NFOG) granted financial support to the planning, data collection and communication of results from this project. The funding sources had no influence on the study design, analysis, interpretation of data, or decision to submit the paper.

## Abstract

**Introduction:** Population-based studies about the consequences of SARS-CoV-2 infection (COVID-19) in pregnancy are few and have limited generalizability to the Nordic population and health care systems.

**Material and method:** This study examines pregnant women with COVID-19 in the five Nordic countries. Pregnant women were included if they were admitted to hospital between March 1 and June 30, 2020 and had a positive SARS-CoV-2 PCR test 14 days or fewer prior to the admission. Cause of admission was classified as obstetric or COVID-19 related.

**Results:** In the study areas, 214 pregnant women with a positive test were admitted to hospital, of which 56 women needed hospital care due to COVID-19. The rate of admission due to COVID-19 was 0.4 per 1000 deliveries in Denmark, Finland, and Norway and 3.8 per 1000 deliveries in the Swedish regions. Women hospitalized because of COVID-19 were more frequently obese (*P* < 0.001) and had migrant background (*P* < 0.001) compared to the total population of women who delivered in 2018. Twelve women (21.4%) needed intensive care. Preterm delivery (n=12, 25%, *P* < 0.001) and cesarean delivery (n=21, 43,8%, *P* < 0.001) were more frequent in women with COVID-19 compared to the women who delivered in 2018. No maternal deaths, stillbirths or neonatal deaths were reported.

**Conclusions:** The risk of admission due to severe COVID-19 disease in pregnancy was low in the Nordic countries. A fifth of the women required intensive care and we observed higher rates of preterm and cesarean deliveries. National public health policies appear to have had an impact on the rates of admission due to severe COVID-19 disease in pregnancy. Nordic collaboration is important in collecting robust data and assessing rare outcomes.

**Key Message:** Risk of hospital admission due to COVID-19 infection among pregnant women was low in the Nordic countries, but varied between the countries, which is most likely related to different national public health strategies.

## Introduction

The World Health Organization declared a global pandemic of coronavirus disease in March 2020 (1). During the H1N1influenza pandemic, pregnant women were particularly vulnerable, resulting in increases of maternal and perinatal mortality among those infected (2-8). The WHO running systematic review and meta-analysis about the effect of COVID-19 in pregnancy identified pre-existing comorbidities, and high maternal age and Body Mass Index (BMI) as risk factors for severe infection (9). Preterm birth rate increased among infected women (9). Recent publications from the US indicate that pregnant women are in a higher risk for serious COVID-19 disease compared to non-pregnant women (10, 11). These studies and the majority of studies in the WHO systematic review were performed in settings with limited generalizability to the Nordic populations and health care systems. With the on-going pandemic, population-based studies with larger case numbers and lower risk of bias are crucial for guiding disease surveillance and health management (12). A few population-based studies assessing the risk and consequences of COVID-19 infection in pregnancy have been published (13-15). However, the inclusion criteria comprised all causes of hospital admission which results in heterogenous study populations.

The population in the Nordic countries is relatively uniform, and health care during pregnancy is provided free of charge. All five countries have medical birth registries with mandatory registrations recording maternal and fetal/neonatal outcomes of all births.

The objective of this study was to describe hospital admissions of pregnant women with COVID-19 in the Nordic countries. We present preliminary, aggregated results including the characteristics and medical risk factors, clinical management and outcomes of pregnant women with COVID-19, focusing primarily on the group of women admitted due to COVID-19, during the first four months of the pandemic in the Nordic countries.

## Material and methods

This study is an ongoing prospective observational study in the Nordic countries and part of the Nordic Obstetric Surveillance Study (NOSS) collaboration. The study includes pregnant women with COVID-19 infection and hospital admission for at least 24 hours. COVID-19 infection was defined as detection of viral RNA on a pharyngeal swab 14 days or fewer prior to hospital admission. Cause of admission was classified as obstetric, eg delivery or obstetric complaints, or COVID-19 related.

Primary outcomes included maternal or neonatal admission to intensive care unit (ICU or NICU), COVID-19 pneumonia, maternal mortality (deaths during pregnancy or within 42 days after the end of pregnancy), preterm delivery (delivery before 37 completed weeks of gestation) and perinatal mortality (stillbirth from 22 completed weeks of gestation and early neonatal deaths). We collected information about sociodemographic risk factors (partner status, migrant background, occupation), pregestational chronic diseases (including cardiac, renal, endocrine, psychiatric, hematologic, and autoimmune disease, cancer and HIV), gestational age (GA) at COVID-19 infection, and clinical care such as induction of labor, and mode of delivery.

Calculation of BMI was based on pregestational weight or first recorded weight in pregnancy. Gestational age was based on estimated date of delivery assessed by ultrasound according to national guidelines, or according to last menstrual period for women without an ultrasound dated pregnancy at the time of COVID-19 infection.

Data were entered into a uniform case report form, adapted in each country.

In Denmark (DK), all obstetric units participated. A clinician in each unit prospectively reported cases to a joint electronic database in EasyTrial (easytrial.net, Denmark). Reminders were sent out to secure data completeness and cases were validated by a retrospective registry linkage, with data obtained from the Danish National Patient Register, the Danish National Service Register and the Danish Microbiology Database every second month. Missing cases identified by validation were entered retrospectively. The Danish study was approved by the Danish Patient Safety Authority (reg. no. 31-1521-252) and the regional Data Protection Agency in Region Zealand (reg. no. REG-022-2020).

In Finland (FI), cases admitted to Helsinki University Hospital were included. Most of the COVID-19 patients in Finland during the study period were inhabitants in the Helsinki region (73% (19)), and fewer than three cases were reported to have been admitted to the other university hospitals by November 2020 (personal correspondence OÄ). Consequently, data collection from Helsinki University Hospital alone is assumed to have included most affected cases. Data were entered into the report form by one primary investigator. The study was approved by Helsinki University Hospital. (reg no. HUS1624/2020)

In Iceland (IS), public health authorities recorded all cases of COVID-19, and information about all pregnant women with COVID-19 was forwarded to the Icelandic Birth Registry. Cases were validated by a retrospective registry linkage with data obtained from the Icelandic National Patient Register and the Icelandic Birth Register. Data were entered into the report form by one primary investigator. The study and data collection were approved by Landspitali University Hospital, the National Bioethics Committee (reg.no.VSNb2020050016/03.0I) and the Icelandic Data Protection Authority (reg.no.20-106).

In Norway (NO), data were prospectively collected by the Medical Birth Registry of Norway (MBRN) at the Norwegian Institute of Public Health (NIPH). All hospitals providing care for pregnant women participated. Reminders were sent out bimonthly. Data were entered using an online form to a safe research server at Service for Sensitive Data (TSD), University of Oslo, contracted by MBRN/NIPH. The data collection was approved by the data protection officer at the NIPH (reg. no. 20_11054) and the study was approved by the Western Regional Ethics Committee (reg. no.125890)

In Sweden (SE), data were included from three institutions -Karolinska University Hospital (KUH), Sahlgrenska University Hospital (SaUH) and Skåne University Hospital (SkUH). These institutions are among the major referral hospitals in the country and account for 22% of annual deliveries in Sweden. Data were retrieved from Hospital Discharge Registers and medical records at each participating obstetric unit and entered into a joint electronic database in REDCap (16, 17) hosted at Lund University. The Swedish study was approved by the Swedish Ethical Review Authority (reg. no. 2020-03012) for SUH and SKH and reg.no. 2020-01499 for KUH. The study was registered in accordance with the Personal Data Act and approved by each Data Protection Officer at every involved institution.

The ethical approvals in DK, IS, NO and KUH exempted the studies from the principle of individual consent. In FI ethical approval is not required in register-based studies. In SaUH and SkUH, women received written information about the study including an opt-out possibility. Data were managed and stored in accordance with national regulations and the General Data Protection Regulation (GDPR).

The study sample size was governed by the disease incidence, so no formal power calculation was performed.

Results are reported both in total for all five countries and restricted to DK, FI, IS and NO to account for potential selection bias in the Swedish data, where only tertiary centers were included. Nominal data are presented as numbers and percentages. Continuous variables are presented as the range of country means when normally distributed and when not normally distributed as range of country medians. Comparison data for all deliveries in 2018, the most recent year for which complete data were available, were collected from the national Medical Birth Registry websites (18-22), and by using unpublished statistics provided by the registries. The total number of deliveries was based on the number of deliveries during the study period in Norway and at the included Swedish sites, and an estimation based on the number of annual deliveries in 2019 in DK, FI and IS. Chi-square tests were used to assess differences in outcome frequencies between the cases and the comparison population.

Data analyses were performed using IBM SPSS statistics 21 (SPSS Inc., Chicago, IL, USA) and STATA 16SE.

## Results

Between March 1 and June 30, 2020, we identified 214 pregnant women who were admitted to hospital for any reason with a positive SARS-CoV-2 PCR test 14 days or fewer prior to hospital admission. No women opted out in Sweden. The rate of hospital admission was 0.8 per 1000 deliveries, ranging from 0.5 to 1.0 per 1000 in DK, FI, IS and NO, and to 18.2 per 1000 in the Swedish regions.

Due to different national and regional testing strategies (Table S1), the study groups in the various countries were heterogeneous. We therefore restricted further analyses to the pregnant women who required hospital admission because of COVID-19 disease, as shown in the flowchart (Figure 1). Characteristics of women admitted for any reason are shown in Table S2.

**Figure 1.**
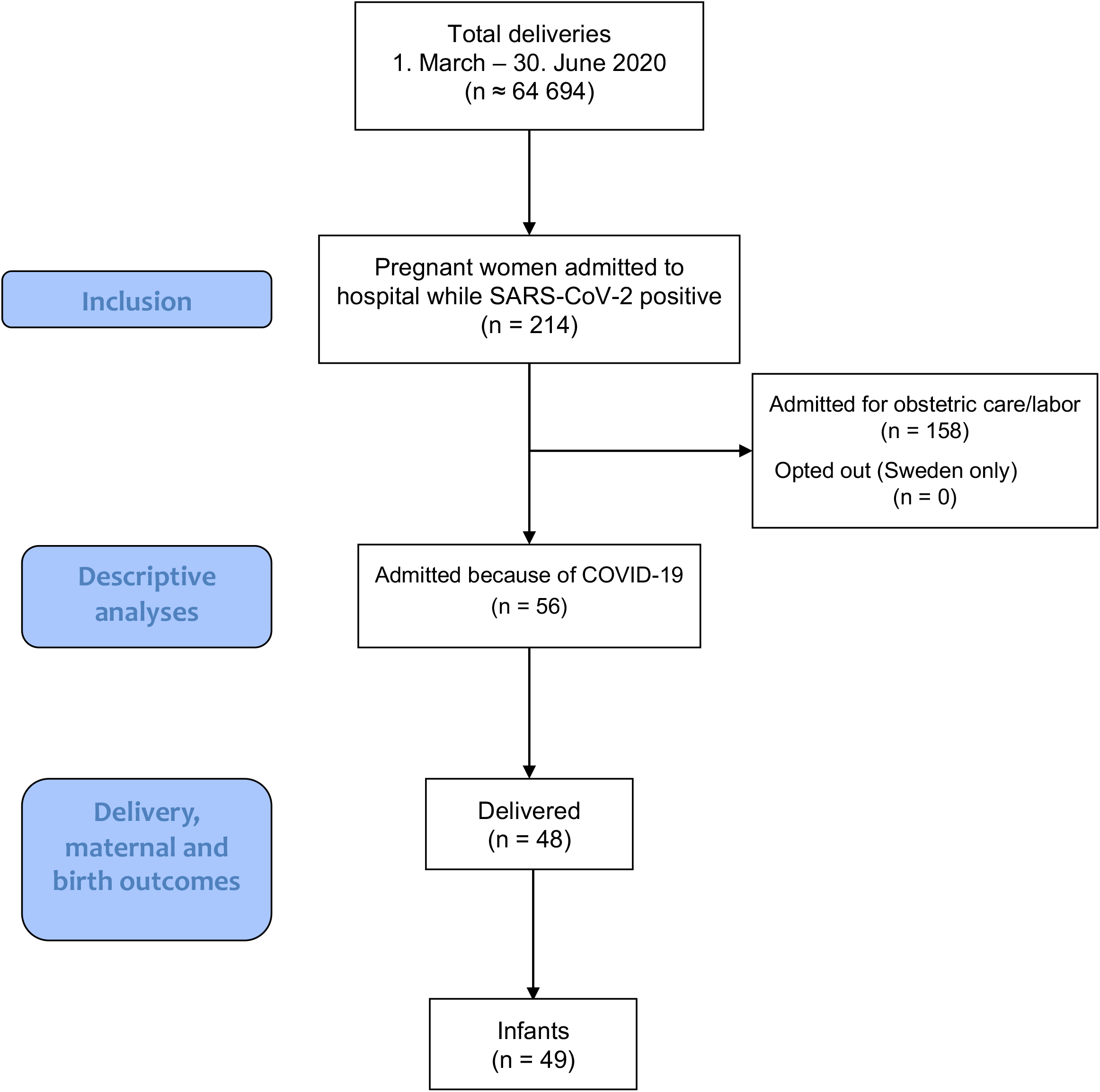
Flow diagram

Fifty-six pregnant women required hospital care for COVID-19 disease. There were no admissions in Iceland and the rate of hospital admission ranged from 0.3 to 0.5 per 1000 deliveries in DK, FI, and NO, and to 3.8 per 1000 in the Swedish regions.

Figure 2 illustrates the gestational age at first positive SARS-CoV-2 test among the included women and Figure S2 illustrates the month of first positive test. Most women admitted due to COVID-19 were in the third trimester of pregnancy when tested positive. Characteristics of the pregnant women with severe COVID-19 requiring hospitalization are presented together with the Medical Birth Registry data from 2018 in Table 1. Compared to the women who delivered in 2018, women admitted due to COVID-19 were more frequently obese, with BMI above 30 (*P* <0.001,) and were migrants (*P* <0.001).

**Table 1.** Characteristics of pregnant women admitted to hospital due to COVID-19 infection in the Nordic countries between March 1 and June 30, 2020 compared to the characteristics of women who delivered in 2018.

|  | NOSS<br>COVID-19<br>population | Deliveries in the<br>Nordic countries<br>2018<sup>a</sup> | P-value<sup>b</sup> | NOSS<br>COVID-19 excluding<br>SE | Deliveries in the Nordic<br>countries excluding SE<br>2018<sup>a</sup> | P-value<sup>b</sup> |
| --- | --- | --- | --- | --- | --- | --- |
| Deliveries<sup>c</sup> | ~ 64 694 | 283 868 |  | ~ 55 255 | 167 779 |  |
| Cases | 56 |  |  | 20 |  |  |
| Hospital admissions due to<br>COVID-19 per 1000 deliveries<br>– estimated risk<sup>d</sup> | ~ 0.9 |  |  | ~ 0.4 |  |  |
| Age,<br>range of means | 29.9 to 33.9 | 30.0 to 31.0 |  | 29.9 to 33.9 | 30.0 to 31.0 |  |
| Age ≥35 years, n (%) | 14 (25.0) | 52 653 (18.6) | 0.219 | 5 (25.0) | 27537 (16.4) | 0.230 |
| BMI,<br>range of means | 24 to 32 | 24,5 to 25,2<sup>e-g</sup> |  | 24 to 32 | 24,5 to 25,2<sup>e-f</sup> |  |
| BMI ≥30, n (%) | 18 (32.1) | 31 509 (14.2)<sup>d-g</sup> | <0.001 | 7 (35.0) | 21 914 (13.3)<sup>d-f</sup> | 0.008 |
| Smoker<sup>i</sup>, n (%) | 2 (3.6) | 16 349 (6.0) | 0.448 | 1 (5.0) | 11 511 (7.3) | 0.697 |
| Immigrant<sup>i</sup>, n (%) | 36 (64.3) | 54 350 (19.2) | <0.001 | 12 (60.0) | 35 206 (21.0) | <0.001 |
| Chronic diseases<sup>k</sup>, n (%) | 16 (28.6) |  |  | 2 (10.0) |  |  |
| Nulliparous, n (%) | 20 (35.7) | 123 416 (43.6) | 0.233 | 7 (35.0) | 74 018 (44.1) | 0.412 |
| Multiple pregnancy, n (%) | 10 (17.9) | 4013 (1.4) | < 0.001 | 1 (5.0) | 2 457 (1.5) | 0.188 |

<sup>a</sup> The Danish Medical Birth Registry, Finnish Medical Birth Registry, Medical Birth Registry of Norway, Icelandic Birth Registry and Swedish Medical Birth Registry annual reports 2018
<sup>b</sup> Chi-square tests.
<sup>c</sup> Estimate based on total deliveries from March-June 2019 in DK, FI, IS<sup>d,e,h</sup>, total deliveries from March-June 2020 in NO<sup>f</sup> and in the SE hospitals.
<sup>d</sup> Danish Medical Birth Register, The Danish Health Data Authority: https://www.esundhed.dk/Emner/Gravide-foedsler-og-boern/Foedte-og-foedsler-1997-#tabpanel61119A72216248AC86DB508579760DED. [accessed 26. november 2020] Mean BMI not available.
<sup>e</sup> Finnish Birth Registry (https://thl.fi/en/web/thlfi-en/statistics/statistics-by-topic/sexual-and-reproductive-health/parturients-deliveries-and-births)
<sup>f</sup> The Medical Birth Registry of Norway, (2018: http://statistikkbank.fhi.no/mfr/, 2020: https://www.fhi.no/hn/helseregistre-og-registre/mfr/svangerskap-og-fodsel-under-koronavirus-pandemien/)
<sup>g</sup> The Swedish Medical Birth Registry 2018 (https://www.socialstyrelsen.se/statistik-och-data/statistik/statistikamnen/graviditeter-forlossningar-och-nyfodda/)
<sup>h</sup> Statistics Iceland: (http://www.hagstofa.is/), Birth Registry data on BMI were not available
<sup>i</sup> Smoking registered in early pregnancy
<sup>j</sup> Women born outside the Nordic countries
<sup>k</sup> Cardiac, renal, endocrine, psychiatric, hematologic, and autoimmune disease, cancer, and HIV
BMI: Body Mass Index (pregestational)

**Figure 2.**
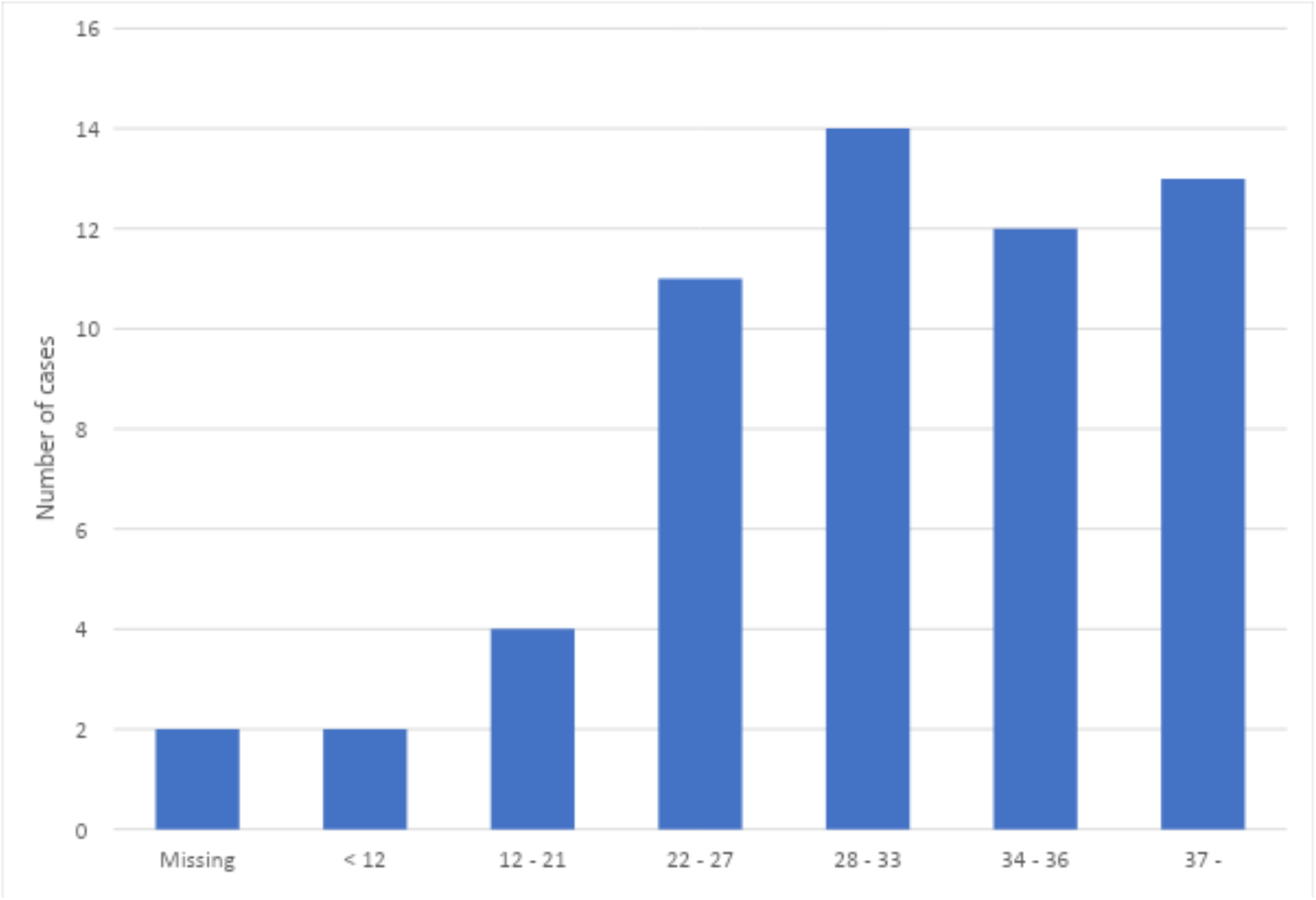
Completed gestational week at first positive SARS-CoV-2 PCR test in 56 pregnant women admitted to hospital due to COVID-19 in the Nordic countries between March 1 and June 30, 2020

In the Swedish regions, relatively more women admitted due to COVID-19 had pre-existing chronic diseases compared to women in the other Nordic countries (SE: N = 14/36; 38.9 %, DK, FI, IS, NO: N = 2/20; 10 %). Multiple pregnancies constituted 28 % (n = 10) of the cases with 90% of these being in Sweden. The Swedish women had positive SARS-CoV-2 tests later in pregnancy and therefore also had a shorter interval from test to delivery.

The maternal and fetal/neonatal outcomes and clinical care are presented in Table 2. Among the 56 cases, 12 women were admitted to an ICU (21.4%). No maternal deaths were reported. During the study period, 48 women delivered 49 infants. Compared to 2018 deliveries, more women admitted due to COVID-19 had a cesarean delivery (41.7 vs 17.3%, *P*<0.001), and the proportion of emergency CD was higher (85.0 vs 53.8%, *P*=0.003). The risk of preterm delivery was also increased (25% vs 5.7%, *P* <0.001). Seven neonates were admitted to NICU (14.3%) and no stillbirths or neonatal deaths were reported.

**Table 2.** Clinical care, delivery, maternal and neonatal outcomes of pregnant women admitted to hospital due to COVID-19 infection in the Nordic countries between March 1 and June 30, 2020 compared to the characteristics of women who delivered in 2018.

|  | NOSS<br>COVID-19<br>population | Deliveries in the<br>Nordic countries<br>2018<sup>a</sup> | *P*-value<sup>b</sup> | NOSS<br>COVID-19<br>excluding SE | Deliveries in the<br>Nordic countries,<br>excluding SE<br>2018<sup>a</sup> | *P*-value<sup>b</sup> |
| --- | --- | --- | --- | --- | --- | --- |
| Total deliveries<sup>c</sup> | ~ 64 694 | 286 868 |  | ~ 55 255 | 167 779 |  |
| Cases | 56 |  |  | 20 |  |  |
| Hospitalization due to COVID-19<br>(per 1000 deliveries) | 0.9 |  |  | 0.4 |  | <0.001 |
| GA at first positive SARS-CoV-2 test, range of<br>country medians | 25w5d to<br>35w2d |  |  | 25w5d to 28w0d |  |  |
| Interval between first positive SARS-CoV-2 test<br>and delivery, days, range of country medians | 31 to 97 |  |  | 68 to 97 |  |  |
| Pneumonia confirmed by imaging, N (%) | 32 (57.1) |  |  | 8 (40.0) |  | 0.188 |
| Admission to ICU, n (%) | 12 (21.4) |  |  | 4 (20.0) |  | 0.893 |
| Maternal death, n (/100.000) | 0 (0) | 6.8 to 8.1d |  | 0 (0) |  |  |
| Delivery outcomes |  |  |  |  |  |  |
| Delivered, n (%) | 48 (85.7) |  |  | 17 (85.0) |  | 0.938 |
| Induction of labor, n (%) | 16 (33.3) | 61592 (21.8) | 0.05 | 5 (29.4) | 43307 (25.8) | 0.735 |
| Mode of delivery |  |  |  |  |  |  |
| Vaginal delivery, n (%) | 28 (58.3) | 233 929 (82.7) | <0.001 | 8 (47.1) | 138 800 (82.7) | <0.001 |
| Cesarean Delivery (CD) | 20 (41.7) | 49 031 (17.3) | <0.001 | 9 (52.9) | 28 979 (17.3) | <0.001 |
| Emergency CD, n (% of all CD) | 17 (85.0) | 26 367 (53.8) | 0.003 | 8 (88.9) | 17 542 (60.5) | 0.082 |
| Elective CD, n (% of all CD) | 3 (15.0) | 22 529 (45.9) | 0.003 | 1 (11.1) | 11 383 (39.3) | 0.084 |
| Preterm delivery GA < 37w, n (%) | 12 (25.0) | 16 211 (5.7) | <0.001 | 5 (29.4) | 9 936 (5.9) | <0.001 |
| Infant outcomes |  |  |  |  |  |  |
| Infants, n | 49 | 286 939 (99.7) | 0.686 | 18 (100) | 169 724 (99.7) | 0.686 |
| Live birth, n (%) | 49 (100) | 286 954 |  | 18 (100) |  |  |
| GA at delivery, range of country medians | 35w4d to 40w1d |  |  | 35w4d to 40w1d |  |  |
| NICU admission, n (%) | 7 (14.3) |  |  | 5 (27.8) |  |  |
| Stillbirth<sup>e</sup> n (/1000) | 0 | 975 (0.34) | 0.686 | 0 | 539 (0.32) | 0.686 |
| Neonatal death<sup>f</sup>, n (/1000) | 0 | 520 (1.8) | 0.768 | 0 | 368 (2.2) | 0.848 |

<sup>a</sup> Danish Medical Birth Register, The Danish Health Data Authority: https://www.esundhed.dk/Emner/Gravide-foedsler-og-boern/Foedte-og-foedsler-1997-#tabpanel61119A72216248AC86DB508579760DED. [accessed 26. november 2020]
Finnish Birth Registry (https://thl.fi/en/web/thlfi-en/statistics/statistics-by-topic/sexual-and-reproductive-health/parturients-deliveries-and-births)
The Medical Birth Registry of Norway, (2018: http://statistikkbank.fhi.no/mfr/, 2020: https://www.fhi.no/hn/helseregistre-og-registre/mfr/svangerskap-og-fodsel-under-koronavirus-pandemien/)
The Swedish Medical Birth Registry 2018 (https://www.socialstyrelsen.se/statistik-och-data/statistik/statistikamnen/graviditeter-forlossningar-och-nyfodda/)
Statistics Iceland: (http://www.hagstofa.is/),
<sup>b</sup> Chi- square tests.
<sup>c</sup> Estimate based on total deliveries from March-June 2019 in DK, FI, IS, total deliveries from March-June 2020 in NO and in the SE hospitals.
<sup>d</sup> Vangen S et al. Maternal deaths in the Nordic countries. Acta Obstet Gynecol Scand 2017; 96:1112–1119.
<sup>e</sup> Stillbirth at gestation age $\geq 22$ weeks or birthweight $\geq 500$ g
<sup>f</sup> Neonatal death within 28 days after birth.

In the Swedish regions, induction of labor among the COVID-19 infected women was more frequent than in the other Nordic countries (SE: N = 11/31; 35.5 %, DK, FI, IS, NO: N = 5/17; 29.4 %) but fewer women had a CD (SE: N = 12/31; 38.7 %, DK, FI, IS, NO: N = 9/17; 52.9 %). In DK, IS, FI, NO more neonates were admitted to NICU compared to in SE (DK, IS, FI, NO: N = 5/18; 27.8%, SE: N = 2/31; 6.5%).

## Discussion

This prospective study in the five Nordic countries showed a low risk of hospital admission of pregnant women with COVID-19 infection. Pregnant women hospitalized because of COVID-19 infection were more often obese and more likely to have a migrant background compared to women who delivered in 2018. Hospitalization due to COVID-19 was associated with an increased risk of delivering preterm and by cesarean.

We included only the most severe cases admitted to hospital due to COVID-19 symptoms in the analyses and excluded women admitted for obstetric reasons, resulting in a study population less influenced by the national testing strategies. These strategies varied over time and between countries during the inclusion period (Figure S1) and could have biased the study results otherwise.

Large population-based cohort studies from the United Kingdom (UK) and Italy found an incidence of hospitalization with COVID-19 of 2/1000 maternities (13-15), which is much higher than the risk of 0.4/1000 deliveries found in DK, FI, IS and NO. However, these studies included admissions for any reason, such as obstetric care and labour. Further, our Nordic study was restricted to women with present infection with a limitation of 14 days between test and admission. Nevertheless, even if all admissions were included in the Nordic countries, the admission risk in DK, FI, SI and NO was lower than in Italy and the UK (0.8/1000 deliveries). Additionally, the rate of admission was considerably higher in the SE regions, which may reflect both higher population infection rates, but also selection bias with higher admission rates at university hospitals. The risk of hospital and intensive care admissions in the total population was also higher in SE compared to the other Nordic countries as illustrated in Figure S3. This may indicate that policies reducing the transmission in the general population also reduces the rate of hospital admission due to COVID-19 among pregnant women.

Among pregnant women admitted with COVID-19 in the Nordic countries, obesity and migrant background were more common than in the 2018 birth population, corresponding to the findings of previous studies (13, 14, 23). This information is relevant when developing national public health strategies.

In the Nordic countries, no maternal, fetal or neonatal deaths were registered among the pregnant women admitted with severe COVID-19 symptoms during the first four months of the pandemic. However, 21% of the women needed intensive care, which is higher than the proportion in previous population-based studies from UK, Italy and the Netherlands (13, 14, 23), reflecting our inclusion of only the most severe cases. Previous Swedish and Dutch studies found that pregnant women with COVID-19 had higher risk of ICU admission compared to non-pregnant women of the same age with COVID-19 (23, 24).

Induction of labor, preterm delivery, CD and emergency CD were more frequent among women with severe COVID-19 requiring hospital admission compared to the 2018 birth population, which corresponds to previous findings (9, 13, 23). An increasing risk of preterm delivery and CD with increasing severity of COVID-19 has also been reported (14). A previous study from one of the including SE centers found no increased risk of severe outcomes (25), however, they included COVID-19 positive women independently of severity, which might have biased their results.

Compared to the other Nordic countries, more women admitted to the included hospitals in SE had other chronic diseases and multiple pregnancies, which possibly reflects selection bias by inclusion of cases from tertiary care hospitals only. In the one of the three centers in SE, they implemented universal testing of all obstetric admissions during the inclusion period (Suppl. Fig. 2). This is reflected in the higher gestational age at first SARS-CoV-2 test and the shorter interval between test and delivery, since relatively more women with scarce symptoms were screened positive upon admission for delivery. Further, fewer women in SE than in the other Nordic countries delivered preterm or had a cesarean delivery, and fewer infants required neonatal care, which reflects that only the most severe cases were identified in the other countries where different testing strategies were applied.

This study has several strengths. Compared to previous studies, the inclusion of a homogeneous group of only severe cases (pregnant women admitted to hospital because of COVID-19 infection and with a positive test within 14 days of admission), strengthens the outcomes and controls for variation in testing strategies between the countries. Further, the data was prospectively recorded in medical records and later retrieved, and all reported cases have been verified with patient records. Additionally, this study assesses the consequences of COVID-19 by having a comparison group not affected by a pandemic. The Nordic collaboration between several countries with relatively uniform populations provides a larger cohort compared to national studies alone, providing relevant data to Nordic and international health care providers.

This study also has several limitations. We only reported outcomes from the most severe cases and cannot therefore draw conclusions about COVID-19 infection among pregnant women in general. Additionally, the aggregate data currently available did not allow for assessment of individual risk factors and mediating factors. Prospective reporting could potentially cause underreporting of cases by lack of identification. Assessment of completeness by linkage to the National Infectious Disease and the Medical Birth Registry was done in DK and Iceland, but was not yet possible in the other countries. However, the number of cases in DK was comparable to that in FI, IS and NO, where testing strategies were similar, indicating that most cases were identified. The Swedish data represent three large tertiary centers, equivalent to 22% of the annual deliveries in SE, and there is a potential risk for both underreporting and selection bias in the SE data. We therefore presented data for all countries combined and for DK, IS, FI and NO alone.

We did not have concurrent data for non-infected pregnancies available for comparison at the time of publication. We therefore relate the results to 2018 data not influenced by the pandemic. The NOSS COVID-19 group plans to analyse data against population data for 2020 when it is released from the Nordic Medical Birth Registries.

Studies similar to the NOSS COVID-19 collaboration are taking place in several countries worldwide as part of the International Network of Obstetric Survey Systems (26). Uniform international population-based studies are needed to reduce potential selection bias in institution-based studies. Combining data internationally will allow assessment of rare, severe complications and risk factors, and aid in the understanding of the disease in pregnancy even better.

## Conclusion

This multi-national Nordic study showed a low risk of admission due to COVID-19 in pregnancy in the Nordic countries. Women admitted to hospital due to COVID-19 were more frequently obese or had migrant background compared to non-infected women. A fifth of the admitted women required intensive care and we observed higher risks of preterm and cesarean deliveries. The study indicates that the risk of admission and complications in pregnancy related to COVID-19 depend on the national public healthcare measures to reduce transmission of disease.

The Nordic collaboration is important in collecting robust data and may provide future benefits in the analysis of rare obstetric outcomes in pandemic responses.

## Data Availability

Additional data will not be available.

## Acknowledgements

The NOSS group thanks professor Marian Knight, DPhil, National Perinatal Epidemiology Unit, Nuffield Department of Population Health, University of Oxford, UK for her initiation of the INOSS study and contribution to planning this Nordic study.

In Denmark we would like to thank the reporting clinicians at the Danish units (in alphabetical order): Eva K Andersen, Charlotte Sander Andersen, Line Strand Andersen, Lise Lotte Torvin Andersen, Charlotte Brix Andersson, Anne-Line Brülle, Lars Burmester, Tine Clausen, Lene Friis Eskildsen, Richard Farlie, Arense Gulbech, Lea Hansen, Lone Hvidman, Mette Holm Ibsen, Fjola Jonsdottir, Lisbeth Jønsson, Mohammed Khalil, åse Klemmensen, Birgitte Lindved, Julie Milbak, Kamilla Gerhard Nielsen, Monica Lund Pedersen, Elisabeth Rønneberg, Morten Beck Sørensen, Anne Nødgaard Sørensen, Manrinda Kaur Tatla, Dorthe Thisted, Annette Thorsen-Meyer, Karen Wøjdeman, Marianne Vestgaard.

In Norway we would like to thank the reporting clinicians:

Katharina Laine, Hilde Christin Lie, Rebecka Dalbye, Kristine Espedal Kaada, Kristin Urnes, Bassam Odicho, Kristin Løvall, Sindre Grindheim, Birte Toft Haugland, Randi Mette Sygard Steen, åse-Turid Rosseland Svoren, Rune Karlsholm Riise, Helena Erlandsson, Cecilie Lysgård, Siri Kojen Andersen, Elisabeth Magnussen, Marit Heggdal, Tina Bjørsvik Eilertsen, Bente Hjelseth, Sølvi Hestnes, Siri Strand Pedersen, Branislav Rosic, Heidi Frostad Sivertsen, Linda Sandsund, Lisa Jakobsen, Marianne Solhaug, Hallvard Fjelltun, Kari Fiske, Nina Høyer, Hilde Bjerkan, Heidi Løntjern, Guro Stokke, Sølvi Lomsdal, Stig Rekkedal Hill, Elin Bolme Haugen, Birgitte Sanda, Karin Lillejord Kristoffersen, Bjørg Else Wallumrød, Marie Therese Hove, Anne Grønnevik.

## Tweetable abstract

Nordic pregnant women with severe COVID-19 requiring hospital admission have increased risk of intensive care unit admission, preterm delivery and cesarean delivery.

## Abbreviations

BMI: Body Mass Index
COVID-19: SARS-CoV-2 disease with symptoms
CD: Cesarean Delivery
DK: Denmark
FI: Finland
GA: Gestational age
ICU: Intensive Care Unit
IS: Iceland
MBRN: the Medical Birth Registry of Norway
NICU: Neonatal Intensive Care unit
NIPH: the Norwegian Institute of Public Health
NO: Norway
SE: Sweden
UK: United Kingdom

## Table and figure legends

**Supplementary Table S1.**
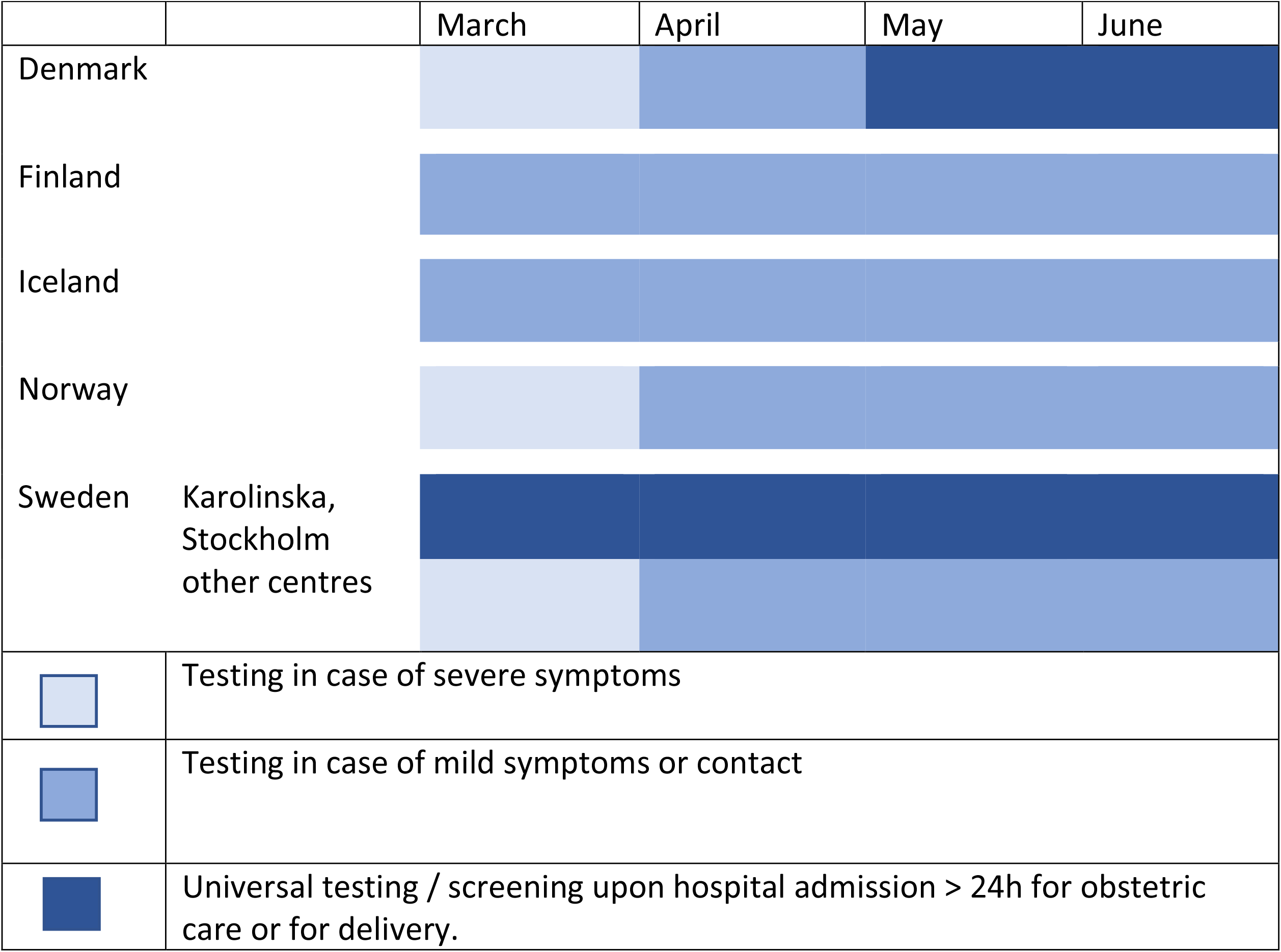
Strategy for COVID-19 testing in each country

**Supplementary Figure S1.**
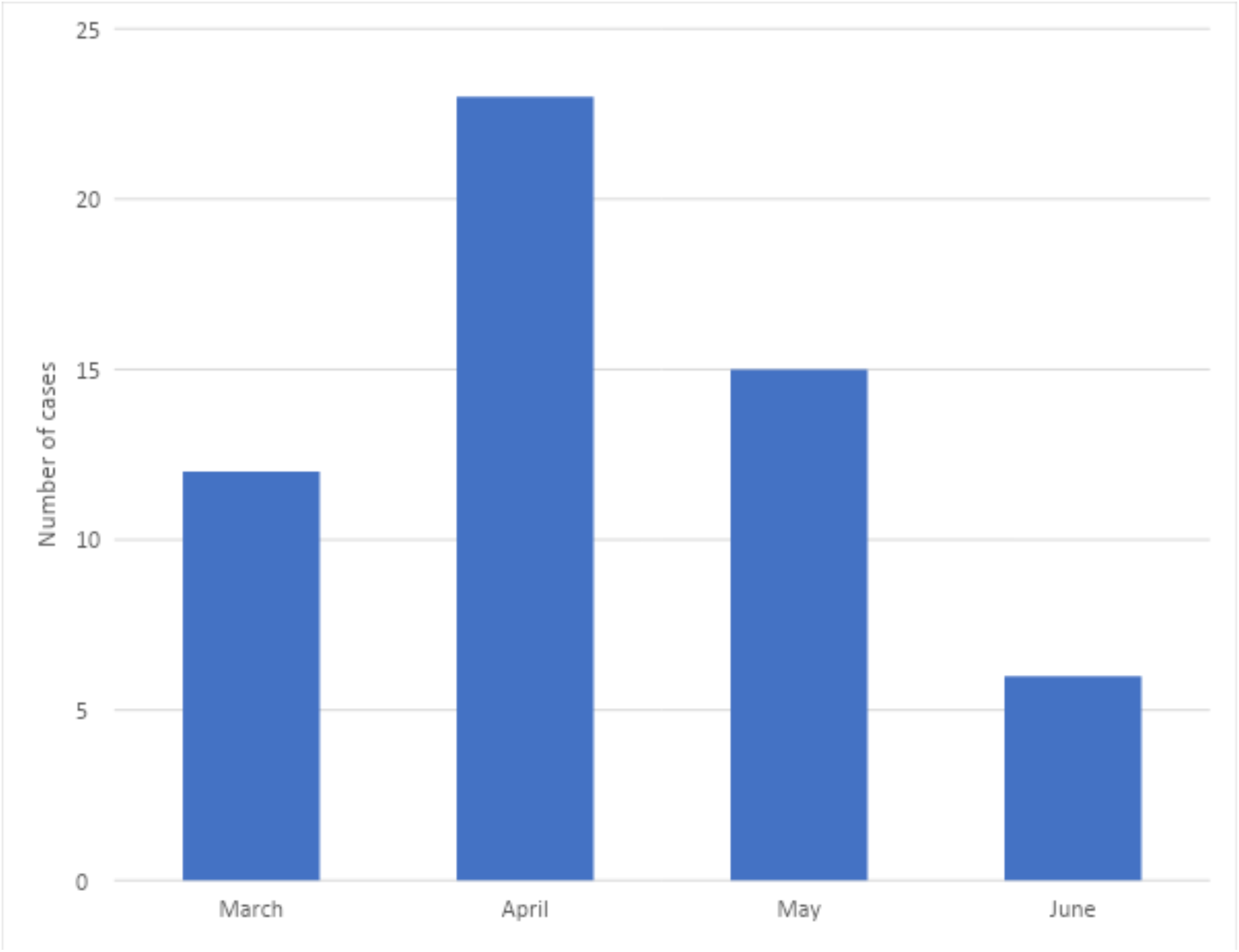
Month of first positive SARS-CoV-2 PCR test in 56 pregnant women admitted to hospital due to COVID-19 in the Nordic countries, March to June 2020

**Supplementary Figure S2.**
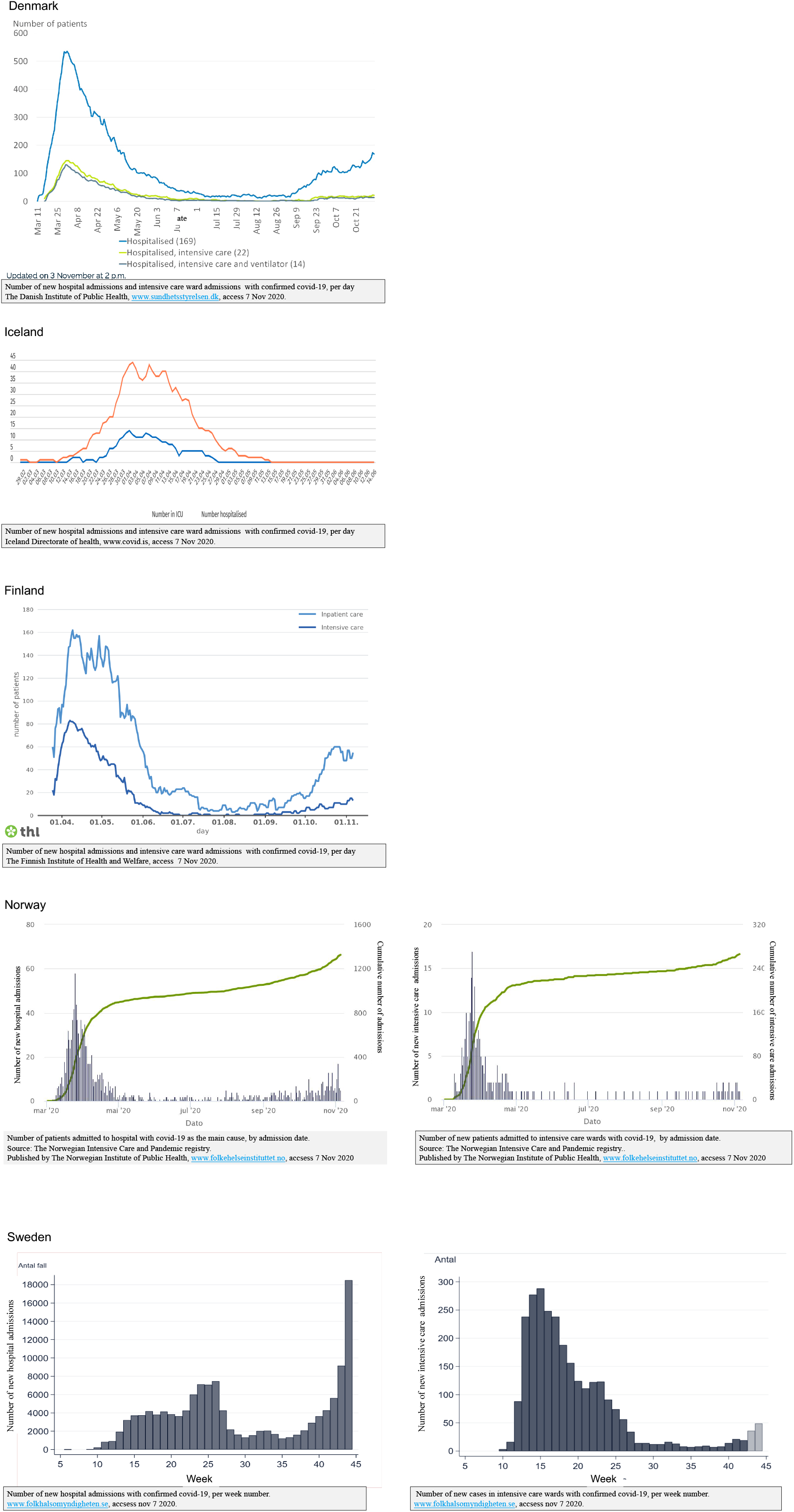
Hospital admissions and intensive care unit admissions due to COVID-19 in the Nordic countries, March to October 2020.

**Supplementary Table S2.** Characteristics of pregnant women admitted to hospital for any reason while having COVID-19 in the Nordic countries between March 1 and June 30, 2020 compared to the characteristics of women who delivered in 2018.

|  | NOSS<br>COVID-19<br>population | NOSS<br>COVID-19 excluding<br>SE | Deliveries in the Nordic<br>countries 2018<sup>a,b</sup> |
| --- | --- | --- | --- |
| Deliveries<sup>a,c</sup> | ~ 64 694 | ~ 55 255 | 282 960 |
| Cases | 214 | 42 |  |
| Hospital admissions with COVID-19 per 1000 deliveries – estimated risk<sup>c</sup> | ~3.3 | ~ 0.8 |  |
| Age,<br>range of means | 31.1 to 33.5 | 31.1 to 33.5 | 30.0 to 31.0 |
| Age ≥35 years, n (%) | 62 (28.9) | 19 (31.0) | 20.3 to 23.6 |
| BMI,<br>range of means | 24 to 33 | 24 to 33 | 24.5 to 25.2<sup>e-g</sup> |
| BMI ≥30, n (%) | 49 (28.4) | 8 (19.0) | 12.7 – 15.5<sup>d-g</sup> |
| Smoker<sup>i</sup>, n (%) | 6 (3.5) | 1 (2.4) | 1.8–5.5 |
| Immigrant<sup>j</sup>, n (%) | 110 (64.0) | 18 (43.0) | 24.5-29.6<sup>f, g</sup> |
| Chronic diseases<sup>k</sup>, n (%) | 66 (38.4) | 4 (9.5) |  |
| Nulliparous, n (%) | 83 (48.3) | 14 (33.0) | 41.9 to 48.3<sup>a</sup> |
| Multiple pregnancy, n (%) | 22 (10.3) | 2 (4.8) | 1.3 to 1.5<sup>a</sup> |

<sup>a</sup> The Danish Medical Birth Registry, Finnish Medical Birth Registry, Medical Birth Registry of Norway, Icelandic Birth Registry and Swedish Medical Birth Registry annual reports 2018
<sup>b</sup> Estimates reported as range of means or range of medians.
<sup>c</sup> Estimate based on total deliveries from March-June 2019 in DK, FI, IS<sup>d,e,h</sup>, total deliveries from March-June 2020 in NO<sup>f</sup> and in the SE hospitals.
<sup>d</sup> Danish Medical Birth Register, The Danish Health Data Authority: https://www.esundhed.dk/Emner/Gravide-foedsler-og-boern/Foedte-og-foedsler-1997-#tabpanel61119A72216248AC86DB508579760DED. [accessed 26. november 2020] Mean BMI not available.
<sup>e</sup> Finnish Birth Registry (https://thl.fi/en/web/thlfi-en/statistics/statistics-by-topic/sexual-and-reproductive-health/parturients-deliveries-and-births)
<sup>f</sup> The Medical Birth Registry of Norway, (2018: http://statistikkbank.fhi.no/mfr/, 2020: https://www.fhi.no/hn/helseregistre-og-registre/mfr/svangerskap-og-fodsel-under-koronavirus-pandemien/)
<sup>g</sup> The Swedish Medical Birth Registry 2018 (https://www.socialstyrelsen.se/statistik-och-data/statistik/statistikamnen/graviditeter-forlossningar-och-nyfodda/)
<sup>h</sup> Statistics Iceland: (http://www.hagstofa.is/), Birth Registry data on BMI were not available
<sup>i</sup> Smoking registered in early pregnancy
<sup>j</sup> Women born outside the Nordic countries
<sup>k</sup> Cardiac, renal, endocrine, psychiatric, hematologic, and autoimmune disease, cancer, and HIV
BMI: Body Mass Index (pregestational)
<sup>b</sup> Chi-square tests.

